# Joint association of genetic risk and accelerometer-based step count with cardiovascular disease: a UK-Biobank cohort study

**DOI:** 10.1101/2025.01.26.25321145

**Authors:** Panagiota Birmpili, Laura Portas, Thomas J. Littlejohns, Aiden Doherty

**Affiliations:** Nuffield Department of Population Health, University of Oxford, Oxford, OX3 7LF, United Kingdom

## Abstract

**Background:** This population-based prospective cohort study aimed to investigate whether accelerometer-measured step count is associated with incident cardiovascular disease (CVD), independently from genetic risk.

**Methods:** The study included participants in the UK Biobank with valid accelerometer and genetic data and without prevalent CVD at baseline. Genetic risk for CVD was categorised as low (1^st^ fifth), moderate (2^nd^-4^th^ fifths), and high (5^th^ fifth). Median daily step count was categorised as low (<6,500), moderate (6,500-12,499), and high (≥12,500). The association of genetic risk and step count with incident CVD, defined as a composite of coronary artery disease and ischaemic stroke, was examined using adjusted Cox proportional hazards models.

**Results:** Of 84,286 participants, 4,847 were diagnosed with CVD during follow-up (median 7.9 years). High genetic risk and low daily step count had a log-additive association with incident CVD. In low genetic risk individuals, step count was not associated with incident CVD. However, in the moderate and high genetic risk groups, those with low step counts had 24% (HR 1.24; 95% Confidence Interval [CI] 1.10-1.40) and 37% (HR 1.37; 95% CI 1.14-1.65) higher risk of incident CVD compared to those with high step counts. There was an inverse dose-response association between the hazard of CVD and step counts up to 10,000 steps/day, which then plateaued in moderate and high genetic risk groups.

**Conclusions:** High daily step count was associated with lower CVD risk in individuals with moderate and high genetic risk, indicating that walking should be encouraged for all, especially those predisposed to CVD.

## Introduction

Cardiovascular disease (CVD) is a major cause of morbidity and mortality globally, with an increasing prevalence due to the aging population, increasing prevalence of obesity and diabetes and changing dietary patterns.^1^ Various modifiable and non-modifiable risk factors have been associated with higher risk of CVD. Multiple genetic variants conferring predisposition to CVD have been found and summarised into polygenic risk scores (PRS), which can be used to identify individuals at higher risk of CVD for primary prevention purposes.^2,3^ Among modifiable risk factors, the role of physical activity has also been well established through large cohort studies, which indicate an inverse dose-response association between physical activity and CVD incidence.^4,5^ Due to the complex gene-environment interactions and their impact on disease development, it is important to consider both genetic predisposition and physical activity on CVD risk.^6^

However, studies investigating the joint association of genetic and lifestyle factors with incident CVD are limited. Most studies rely on self-reported physical activity,^7–13^ which is prone to recall and social desirability bias.^14,15^ Recent studies using wearable devices, which measure physical activity levels more accurately than self-report, have investigated activity intensity levels, such as light, moderate or vigorous, which are often difficult to interpret or use to develop guidance for the public.^16,17^

A more intuitive measure of physical activity is the number of steps a person takes daily, which is readily available on mobile phones and commercial wearable devices. There is accumulating evidence for a non-linear inverse dose-response association between step counts and cardiovascular outcomes, which could be translated into actionable public health policies recommending a minimum number of steps per day for optimal health.^18,19^ However, no studies have explored whether objectively measured step count can modify the association between genetic risk and cardiovascular incidence. We therefore aimed to investigate whether accelerometer-measured step count was associated with the incidence of CVD independently from genetic risk.

## Methods

### Study population

The UK Biobank (UKB) is a prospective cohort of over 500,000 individuals aged 40-69 years living in England, Scotland, and Wales at the time of recruitment, from 2006 to 2010.^20^ The UK Biobank received ethical approval from the North West Multi-centre Research Ethics Committee (16/NW/0274), and all participants provided written informed consent before taking part. Upon enrolment in the study, participants completed a touchscreen questionnaire, underwent verbal interviews, had physical measurements, and provided blood samples for biochemical and genetic analysis. From 1^st^ June 2013 to 31^st^ December 2015, participants with an email address were invited to participate in an accelerometer study, during which they were required to wear an Axivity AX3 triaxial accelerometer on their dominant wrist for seven days.^21^ Participants were excluded from our study if they had missing or poor-quality accelerometer data, defined as less than 72 hours of wear time, lack of data in each one-hour period of the 24-hour cycle, device calibration errors, implausibly high average acceleration (>100 milligravity units), unrealistically low median daily step count (<50 steps/day), or missing cadence data.^22^ Participants with missing values on the exposures and covariates of interest and those with existing cardiovascular disease at the time of the accelerometer wear based on self-reported information or hospital records (Supplementary Table 1) were also excluded.

### Genetic risk

The standard CVD PRS in the UK Biobank was used as a marker of genetic susceptibility to CVD, and its development is described in detail elsewhere.^23^ In summary, UK Biobank participants were genotyped using a custom Axiom array of 825,927 genetic variants, followed by genome-wide imputation to approximately 96 million variants using the Haplotype Reference Consortium and the UK10k/1000 Genomes reference panels. The standard PRS for CVD was then constructed by performing fixed-effect inverse variance meta-analysis of external summary statistics from the three largest genome-wide association studies of CVD.^23^ We used a raw PRS value that was calculated for each UKB participant as the genome-wide sum of the per-variant posterior effect size multiplied by allele dosage, and then a corrected PRS value was generated by centering and variance-standardising the PRS to achieve a standard normal distribution with zero mean and unit variance, while accounting for an individual’s inferred genetic ancestry group.^23^ The PRS distribution was divided into fifths, categorising participants into low (1^st^ fifth), moderate (2^nd^-4^th^ fifths), or high (5^th^ fifth) genetic risk groups, similar to other studies.^7,24^

### Median daily step count

To measure step count from raw accelerometer data, a hybrid self-supervised machine learning and peak detection algorithm was used, where an activity classification model first detected periods of walking and non-walking, followed by step counting only on predicted walking data periods.^22^ This was validated against reference video measurements. Missing step count data from non-wear was imputed by averaging the step count from the corresponding time of day in all other valid days. Overall daily step count was reported as median number of steps taken across the seven-day measurement window and was used as marker of physical activity. It was explored as a categorical variable, divided into fifths and then categorised into three groups: low (1^st^ fifth; <6,500 steps/day), moderate (2^nd^-4^th^ fifth; 6,500-12,499 steps/day), and high (5^th^ fifth; ≥12,500 steps/day). The cut-off points were rounded to the nearest 500 steps to aid with interpretability. The median daily step count was also explored as a continuous variable using restricted cubic spline models with knots placed at the 10th, 50th, and 90th percentiles of the step count distribution, as the association with CVD was not linear.

Peak-30 cadence, defined as the average steps per minute for the 30 highest, but not necessarily consecutive minutes per day, was used as a secondary marker of accelerometer-measured physical activity. It was also divided into fifths and then categorised into three groups (low-1^st^ fifth, moderate-2^nd^-4^th^ fifths, high-5^th^ fifth). While step count provided information on the quantity of physical activity, peak cadence represented an individual’s walking pace, adding the dimension of activity intensity.

To examine the joint association of genetic risk and step count, a variable was constructed by combining step count and genetic risk categories, using the group with low genetic risk and high step count (ideal scenario) as reference, in keeping with other studies.^7,25^ Another combined variable was constructed from the cadence and genetic risk categories, with low risk-high cadence being the reference category.

### Covariates

Known risk factors for CVD, such as demographic characteristics, lifestyle factors, and comorbidities, were used as covariates in this study.^1^ Demographic variables included age at the time of accelerometer wear, sex (male, female), ethnicity (white, non-white), educational attainment (basic education, further education, university degree), employment status (employed, not employed, retired), country of residence (England, Scotland, Wales), and area-level socioeconomic deprivation represented by the Townsend Deprivation Index (TDI) divided into fifths based on the national TDI distribution. Sex was acquired from electronic patient records at recruitment and occasionally updated by the participant, with the options being male or female. Lifestyle risk factors included smoking status (never, previous, current smoker), alcohol consumption (daily, ≥1-4 times/week, <1 time/week, never), accelerometer-measured average sleep duration per night (≤6h, 7–8h, ≥9h),^26^ and diet. Aspects of diet captured through self-reported questionnaires were fruit and vegetable consumption (<5, 5-7.9, ≥8 servings/day), oily and non-oily fish consumption (<2, ≥2 times/week), red or processed meat consumption (<1, 1-3, 3-4, ≥5 times/week), and salt intake (never/rarely, sometimes, usually/always). The family history of CVD referred to self-reported cardiovascular conditions of the father, mother, or siblings. Diabetes mellitus was identified through self-reported diagnosis at baseline, use of medications for diabetes, random glucose levels of ≥11.1 mmol/L or haemoglobin A1c levels >42mmol/mol. Hypertension was ascertained through self-reporting in the baseline questionnaire or interview, use of antihypertensive medications, measured mean systolic blood pressure (SBP) of ≥140mmHg or mean diastolic blood pressure (DBP) of ≥90mmHg. SBP and DBP were calculated as the mean of consecutive manual or automatic measurements at baseline and were explored as continuous variables. The presence of hypercholesterolaemia was captured through self-reported diagnosis, use of lipid-lowering medications, or a total cholesterol of ≥5 mmol/L. Body mass index at baseline was calculated from height and weight measurements in kg/m^2^ and was entered in the models as continuous variable. Finally, the season of accelerometer wear was also included in the models, because the levels of activity were higher in spring and summer months compared to autumn and winter. Information about these variables is provided in Supplementary Table 2.

### Outcomes

The primary outcome was the incidence of non-fatal and fatal CVD, defined as a composite of coronary artery disease (CAD) and ischaemic stroke (IS), ascertained through linkage with electronic hospital inpatient records and mortality registers using a combination of diagnosis and procedure codes. The International Classification of Diseases (ICD-9 and ICD-10) was used for coding of hospital diagnoses and causes of death (Supplementary Table 1), while the UK OPCS Classification of Interventions and Procedures (OPCS-4) was used for surgical operations. The follow-up period started at the end of each individual’s 7-day accelerometer wear period and ended on 31^st^ October 2022 for participants in England, on 31^st^ August 2022 for those in Scotland, and on 31^st^ May 2022 for those in Wales. Participants were censored at the time of first CVD event, death, loss to follow-up, or at the end of the follow-up period, whichever occurred first.

### Statistical analysis

Summary statistics were presented as frequencies and proportions for categorical variables, mean and standard deviation for continuous normally distributed variables, and median and interquartile range (IQR) for continuous not normally distributed variables. Baseline characteristics were compared between PRS groups and separately between step count groups using the chi-square test of association for categorical and the F-test from a linear regression model for continuous variables. Univariable and multivariable Cox proportional hazards models were fitted to evaluate the association of step count and PRS with the incidence of CVD, using participants’ age rather than time on study as the underlying timescale.^27^ Multivariable models were adjusted for the above described covariates. The results were reported as log-relative hazard ratios (HRs) and 95% confidence intervals (CIs). The proportional hazards assumption was checked graphically using Schoenfeld residuals tests and log-log plots and was not violated for any of the variables. Analyses were also stratified by genetic risk group, to assess the association of CVD hazard with step count within each group separately by allowing the baseline hazard function to vary between strata. The statistical significance of the interaction term between PRS and step count was investigated using the likelihood ratio test.

Three sensitivity analyses were conducted. The first excluded individuals with less than two and less than four years of follow-up to assess potential reverse causality, as these individuals could have subclinical CVD at the time of the accelerometer study that affected their step counts. A second was performed excluding people with chronic conditions that may have limited their ability to walk at the time of accelerometer wear, such as cancer, cardiovascular diseases other than CAD and IS, chronic lower respiratory diseases, neurological conditions, musculoskeletal and connective tissue disorders, and abnormalities of gait and mobility ascertained from inpatient hospital records, and those who reported long-standing illness or disability during the baseline interview (Supplementary Table 1). A further sensitivity analysis explored the associations of genetic risk and step count with the two components of CVD, CAD and IS, separately.

All statistical tests were two-sided and a p-value of <0.05 was considered statistically significant. Datasets were processed in R version 4.3.1 and statistical analyses were performed in STATA 18.0. The study is reported in accordance with the Strengthening the Reporting of Observational Studies in Epidemiology (STROBE) guidelines.^28^

## Results

Of the 103,660 participants with accelerometer data, 7,663 were excluded due to poor accelerometer data quality, 2,326 due to lack of genetic data, 6,789 due to prevalent CVD and 2,596 due to missing data in covariates, leaving 84,286 individuals in the analysis (Figure 1). Median age at the time of accelerometer wear was 63 years (IQR 56-68), 58.1% (n=48,994) of participants were women and 97.1% (n=81,826) were of white ethnicity. Participants had high socioeconomic status, with high proportion of university educated (44.6%, n=37,563) and employed individuals (63.5%, n=53,562) and low levels of deprivation (least deprived 50.9%, n=42,929). There were significant differences across step count groups for all baseline characteristics (Table 1). Participants in the low step count group were older, less well educated, consumed more red meat and salt and less fish, fruit, and vegetables, and there was higher proportion of smokers, people with diabetes, hypercholesterolaemia, and hypertension, compared to the high step count group. The median daily step count was 9,134 steps (IQR 6,910-11,749) and the mean peak 30-minute cadence was 93.0 (SD 16.3) steps/minute, with no significant differences between genetic risk groups (p=0.099 and p=0.226 respectively) (Supplementary Table 3).

**Figure 1.**
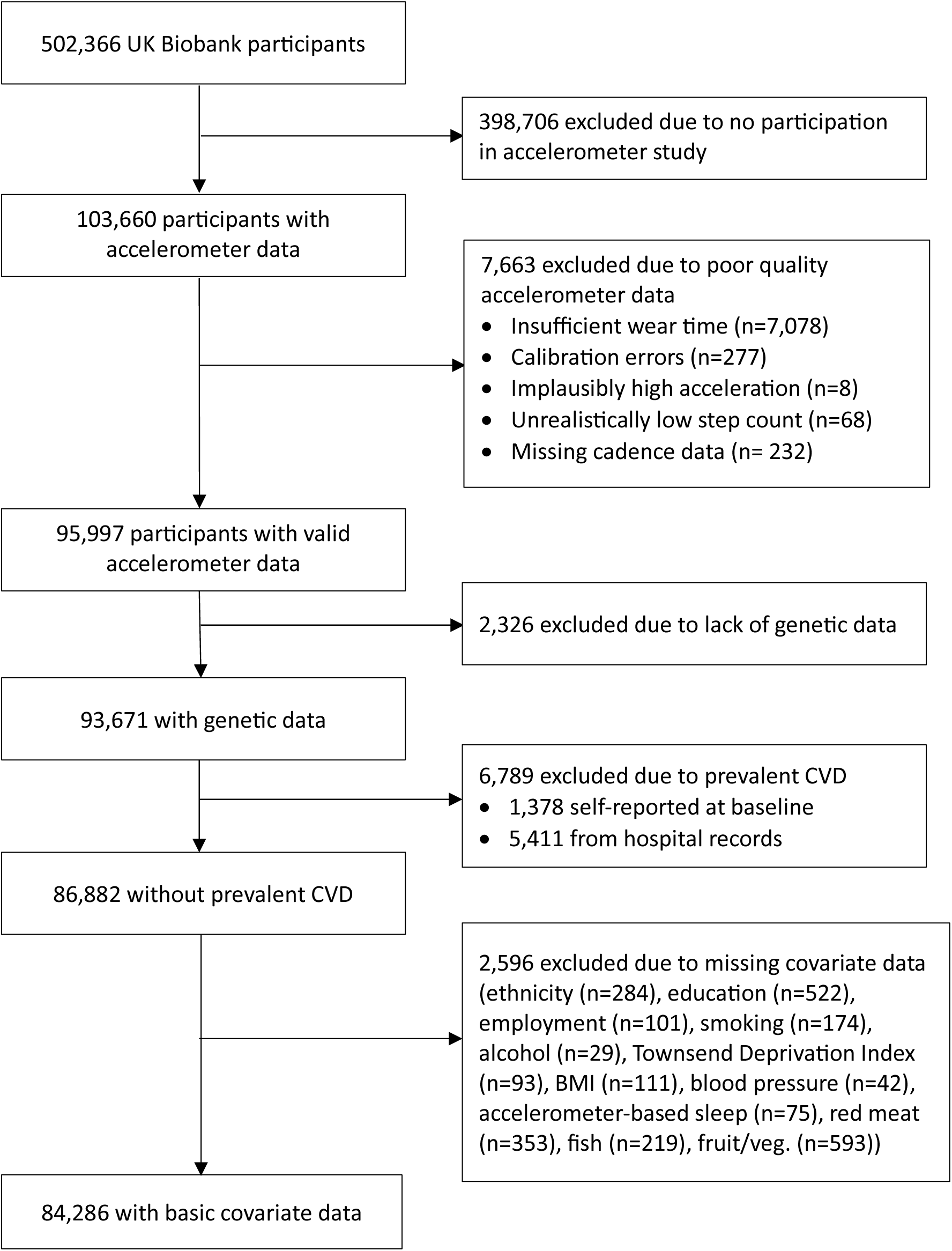
Study participant flow diagram

**Table 1.** Baseline characteristics of study participants by step count group.

|  | Median daily step count |  |  | Total<br>(N=84,286) | p-value |
| --- | --- | --- | --- | --- | --- |
|  | Low<br>(N=17,667) | Moderate<br>(N=49,948) | High<br>(N=16,671) |  |  |
| **Genetic risk** |  |  |  |  | 0.023 |
| Low | 3,401 (19.3) | 10,074 (20.2) | 3,383 (20.3) | 16,858 (20.0) |  |
| Moderate | 10,646 (60.3) | 29,885 (59.8) | 10,040 (60.2) | 50,571 (60.0) |  |
| High | 3,620 (20.5) | 9,989 (20.0) | 3,248 (19.5) | 16,857 (20.0) |  |
| **Age (years)** | 64.0 (56.5-69.2) | 63.0 (55.8-68.2) | 61.9 (55.1-67.2) | 62.9 (55.8-68.2) | <0.001 |
| **Reported sex** |  |  |  |  | <0.001 |
| Female | 10,483 (59.3) | 29,332 (58.7) | 9,179 (55.1) | 48,994 (58.1) |  |
| Male | 7,184 (40.7) | 20,616 (41.3) | 7,492 (44.9) | 35,292 (41.9) |  |
| **Ethnicity** |  |  |  |  | <0.001 |
| White | 17,079 (96.7) | 48,526 (97.2) | 16,221 (97.3) | 81,826 (97.1) |  |
| Nonwhite | 588 (3.3) | 1,422 (2.8) | 450 (2.7) | 2,460 (2.9) |  |
| **Educational attainment** |  |  |  |  | <0.001 |
| Basic education | 4,600 (26.0) | 10,929 (21.9) | 3,213 (19.3) | 18,742 (22.2) |  |
| Further education | 6,345 (35.9) | 16,279 (32.6) | 5,357 (32.1) | 27,981 (33.2) |  |
| University | 6,722 (38.0) | 22,740 (45.5) | 8,101 (48.6) | 37,563 (44.6) |  |
| **Employment status** |  |  |  |  | <0.001 |
| Employed | 10,368 (58.7) | 32,083 (64.2) | 11,111 (66.6) | 53,562 (63.5) |  |
| Not employed | 1,348 (7.6) | 3,011 (6.0) | 1,295 (7.8) | 5,654 (6.7) |  |
| Retired | 5,951 (33.7) | 14,854 (29.7) | 4,265 (25.6) | 25,070 (29.7) |  |
| **Townsend Depriv. Index** |  |  |  |  | <0.001 |
| Least deprived | 8,673 (49.1) | 25,988 (52.0) | 8,268 (49.6) | 42,929 (50.9) |  |
| 2nd Quintile | 4,064 (23.0) | 11,244 (22.5) | 3,714 (22.3) | 19,022 (22.6) |  |
| 3rd Quintile | 2,453 (13.9) | 6,781 (13.6) | 2,456 (14.7) | 11,690 (13.9) |  |
| 4th Quintile | 1,798 (10.2) | 4,457 (8.9) | 1,681 (10.1) | 7,936 (9.4) |  |
| Most deprived | 679 (3.8) | 1,478 (3.0) | 552 (3.3) | 2,709 (3.2) |  |
| **Country** |  |  |  |  | <0.001 |
| England | 15,854 (89.7) | 44,878 (89.8) | 14,886 (89.3) | 75,618 (89.7) |  |
| Scotland | 1,068 (6.0) | 3,175 (6.4) | 1,228 (7.4) | 5,471 (6.5) |  |
| Wales | 745 (4.2) | 1,895 (3.8) | 557 (3.3) | 3,197 (3.8) |  |
| **Smoking status** |  |  |  |  | <0.001 |
| Never | 9,698 (54.9) | 29,542 (59.1) | 9,820 (58.9) | 49,060 (58.2) |  |
| Previous | 6,448 (36.5) | 17,223 (34.5) | 5,887 (35.3) | 29,558 (35.1) |  |
| Current | 1,521 (8.6) | 3,183 (6.4) | 964 (5.8) | 5,668 (6.7) |  |
| **Alcohol consumption** |  |  |  |  | <0.001 |
| Daily | 3,459 (19.6) | 11,657 (23.3) | 4,256 (25.5) | 19,372 (23.0) |  |
| 1-4 /week | 8,418 (47.6) | 26,218 (52.5) | 8,796 (52.8) | 43,432 (51.5) |  |
| < 1/week | 4,516 (25.6) | 9,623 (19.3) | 2,823 (16.9) | 16,962 (20.1) |  |
| Never | 1,274 (7.2) | 2,450 (4.9) | 796 (4.8) | 4,520 (5.4) |  |
| **Red/processed meat** |  |  |  |  | <0.001 |
| <1 time/week | 1,198 (6.8) | 3,918 (7.8) | 1,656 (9.9) | 6,772 (8.0) |  |
| 1-3 times/week | 6,646 (37.6) | 19,531 (39.1) | 6,393 (38.3) | 32,570 (38.6) |  |
| 3-4 times/week | 5,311 (30.1) | 14,672 (29.4) | 4,706 (28.2) | 24,689 (29.3) |  |
| 5+ times/week | 4,512 (25.5) | 11,827 (23.7) | 3,916 (23.5) | 20,255 (24.0) |  |
| **Fish consumption** |  |  |  |  | <0.001 |
| 0-1 times/week | 8,843 (50.1) | 24,128 (48.3) | 8,117 (48.7) | 41,088 (48.7) |  |
| 2+ times/week | 8,824 (49.9) | 25,820 (51.7) | 8,554 (51.3) | 43,198 (51.3) |  |
| **Fruit/veg consumption** |  |  |  |  | <0.001 |
| <5 servings/day | 4,328 (24.5) | 9,891 (19.8) | 3,016 (18.1) | 17,235 (20.4) |  |
| 5-7.9 servings/day | 7,198 (40.7) | 21,492 (43.0) | 7,000 (42.0) | 35,690 (42.3) |  |
| 8+ servings/day | 6,141 (34.8) | 18,565 (37.2) | 6,655 (39.9) | 31,361 (37.2) |  |
| **Salt consumption** |  |  |  |  | <0.001 |
| Never/rarely | 10,304 (58.3) | 30,209 (60.5) | 10,196 (61.2) | 50,709 (60.2) |  |
| Sometimes | 4,697 (26.6) | 13,255 (26.5) | 4,371 (26.2) | 22,323 (26.5) |  |
| Usually/Always | 2,666 (15.1) | 6,484 (13.0) | 2,104 (12.6) | 11,254 (13.4) |  |
| **Sleep duration** |  |  |  |  | <0.001 |
| 6h or fewer | 5,848 (33.1) | 16,492 (33.0) | 6,323 (37.9) | 28,663 (34.0) |  |
| 7-8h | 11,107 (62.9) | 32,348 (64.8) | 10,148 (60.9) | 53,603 (63.6) |  |
| 9h or more | 712 (4.0) | 1,108 (2.2) | 200 (1.2) | 2,020 (2.4) |  |
| **Family history of CVD** | 10,161 (57.5) | 28,027 (56.1) | 9,122 (54.7) | 47,310 (56.1) | <0.001 |
| **Diabetes** | 1,520 (8.6) | 2,441 (4.9) | 665 (4.0) | 4,626 (5.5) | <0.001 |
| **Hypercholesterolaemia** | 14,273 (80.8) | 39,655 (79.4) | 12,983 (77.9) | 66,911 (79.4) | <0.001 |
| **Hypertension** | 9,923 (56.2) | 23,797 (47.6) | 7,372 (44.2) | 41,092 (48.8) | <0.001 |
| **Body Mass Index (kg/m<sup>2</sup>)** | 27 (24-31) | 26 (23-29) | 25 (23-28) | 26 (24-29) | <0.001 |
| **Systolic blood pressure (mmHg)** | 138.0 ± 18.3 | 135.9 ± 18.0 | 135.1 ± 17.9 | 136.2 ± 18.1 | <0.001 |
| **Diastolic blood pressure (mmHg)** | 82.8 ± 10.0 | 81.5 ± 9.9 | 81.0 ± 9.9 | 81.7 ± 10.0 | <0.001 |
| **Median daily steps** | 5,233<br>(4,254-5,918) | 9,189<br>(7,888-10,619) | 14,543<br>(13,378-16,439) | 9,134<br>(6,910-11,749) | <0.001 |
| **Peak 30-min cadence (steps/min)** | 75.1 ± 13.1 | 94.6 ± 12.5 | 107.3 ± 11.9 | 93.0 ± 16.3 | <0.001 |
| **Season of wear** |  |  |  |  | <0.001 |
| Autumn | 5,338 (30.2) | 14,796 (29.6) | 4,742 (28.4) | 24,876 (29.5) |  |
| Winter | 4,529 (25.6) | 10,409 (20.8) | 2,769 (16.6) | 17,707 (21.0) |  |
| Spring | 3,863 (21.9) | 11,479 (23.0) | 4,058 (24.3) | 19,400 (23.0) |  |
| Summer | 3,937 (22.3) | 13,264 (26.6) | 5,102 (30.6) | 22,303 (26.5) |  |

Summary statistics are presented as n (%) for categorical variables, mean ± SD for continuous normally distributed, and median (IQR) for not normally distributed variables. p-values derive
*from Pearson's chi-squared test for categorical variables and linear regression for continuous variables.*

### Association of PRS and step count categories with incident CVD

During a median follow-up of 7.9 years (IQR 7.3-8.4) (total = 644,621 person-years), 4,847 participants (5.8%) experienced a cardiovascular event, 3,746 of which were CAD events and 1,101 ischaemic stroke events. Median daily step count was independently associated with CVD incidence. Compared to the high step count group, those with moderate and low step count had 11% (HR 1.11; 95% CI 1.02-1.20) and 24% (HR 1.24; 95% CI 1.13-1.36) higher hazard of CVD respectively, in fully adjusted models (Table 2). Genetic risk had a stronger independent association with incident CVD than step count, with those in the moderate risk group having 52% (HR 1.52; 95% CI 1.40-1.66) and those in the high risk group having 103% (HR 2.03; 95% CI 1.84-2.23) higher hazard of CVD compared to the low genetic risk group (Table 2).

**Table 2.** Association of genetic risk and median daily step count with incident cardiovascular disease.

|  | Events | Total | Hazard Ratio (95% CI) |  |
| --- | --- | --- | --- | --- |
|  |  |  | Univariable model | Fully adjusted model |
| Genetic risk |  |  |  |  |
| Low | 634 | 16,858 | 1.00 | 1.00 |
| Moderate | 2,930 | 50,571 | 1.60 (1.47-1.74) | 1.52 (1.40-1.66) |
| High | 1,283 | 16,857 | 2.19 (1.99-2.41) | 2.03 (1.84-2.23) |
| P (association) |  |  | <0.001 | <0.001 |
| Median daily step count |  |  |  |  |
| Low | 644 | 7,570 | 1.44 (1.32-1.58) | 1.24 (1.13-1.36) |
| Moderate | 2,517 | 42,165 | 1.12 (1.04-1.22) | 1.11 (1.02-1.20) |
| High | 330 | 6,879 | 1.00 | 1.00 |
| P (association) |  |  | <0.001 | <0.001 |
| Combined variable |  |  |  |  |
| Low genetic risk |  |  |  |  |
| High step count | 116 | 3,383 | 1.00 | 1.00 |
| Moderate step count | 365 | 10,074 | 1.01 (0.81-1.24) | 0.98 (0.80-1.21) |
| Low step count | 153 | 3,401 | 1.19 (0.94-1.52) | 1.04 (0.81-1.32) |
| Moderate genetic risk |  |  |  |  |
| High step count | 471 | 10,040 | 1.42 (1.16-1.74) | 1.35 (1.10-1.65) |
| Moderate step count | 1677 | 29,885 | 1.61 (1.33-1.94) | 1.51 (1.25-1.82) |
| Low step count | 782 | 10,646 | 2.03 (1.67-2.46) | 1.67 (1.37-2.03) |
| High genetic risk |  |  |  |  |
| High step count | 191 | 3,248 | 1.86 (1.48-2.35) | 1.72 (1.36-2.17) |
| Moderate step count | 725 | 9,989 | 2.16 (1.78-2.63) | 1.99 (1.63-2.42) |
| Low step count | 367 | 3,620 | 2.94 (2.39-3.63) | 2.34 (1.89-2.89) |
| P (association) |  |  | <0.001 | <0.001 |

*Median daily step count categories: low (<6,500 steps/day), moderate (6,500-12,499 steps/day), high (≥12,500 steps/day). Genetic risk categories: low (1<sup>st</sup> fifth), moderate (2<sup>nd</sup> – 4<sup>th</sup> fifths), high (5<sup>th</sup> fifth)*

High genetic risk and low daily step count had a log-additive association with higher hazard of CVD, but there was no multiplicative interaction effect (p=0.490) and this term was therefore not included in the models. Compared to the low genetic risk and high step count group, low step count was not associated with higher hazard of CVD in the low genetic risk group (HR 1.04; 95% CI 0.81-1.32), but was associated with 67% (HR 1.67; 1.37-2.03) and 134% (HR 2.34; 1.89-2.89) higher hazard of CVD in the moderate and the high genetic risk groups respectively, after full adjustment (Table 2, Figure 2).

**Figure 2.**
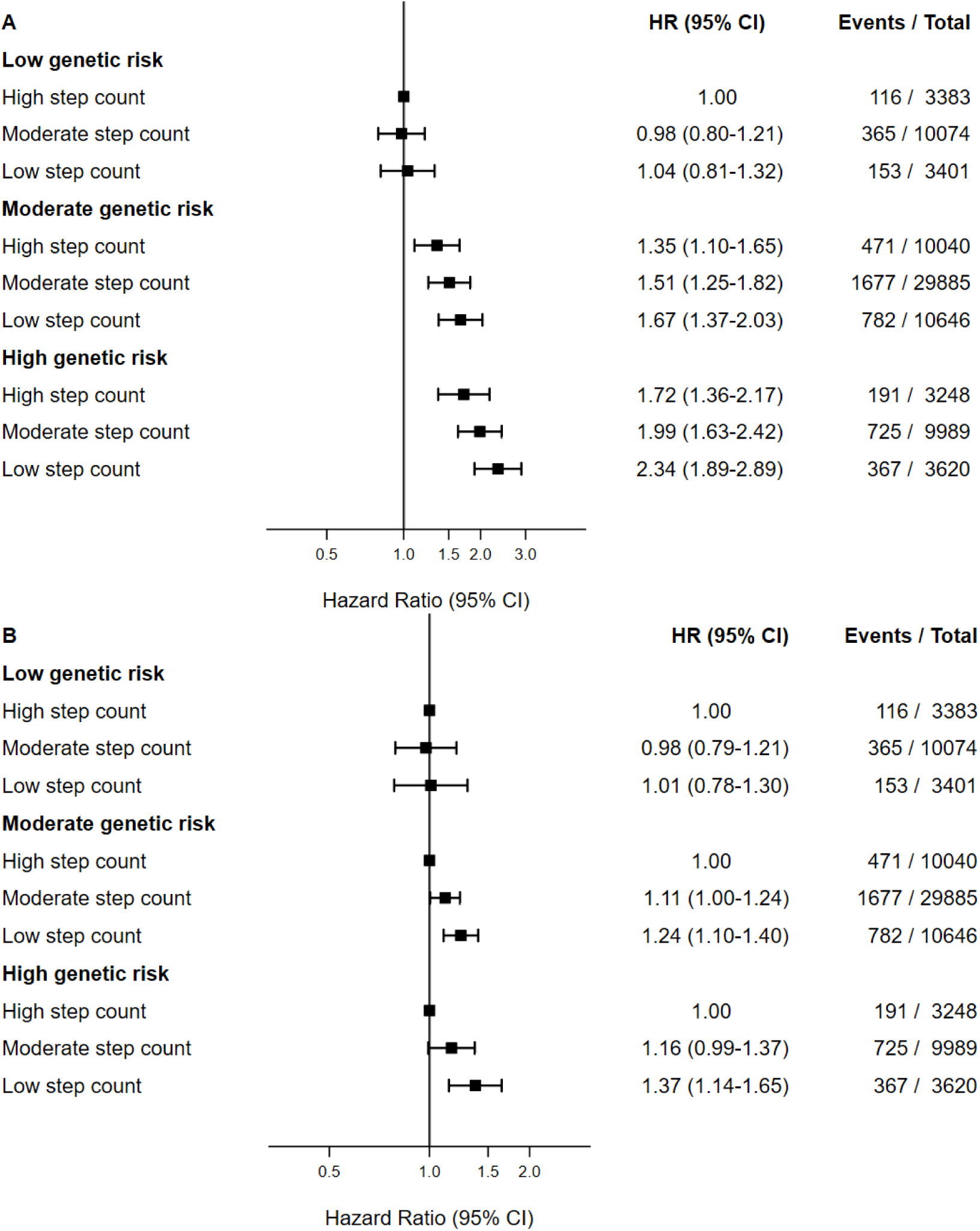
Association of genetic risk and step count with incident cardiovascular disease overall (A) and stratified by genetic risk (B). Squares represent hazard ratios (HR) and the vertical lines 95% confidence intervals

Similar results were found when the association of step count with CVD was explored within each genetic risk group separately in stratified models (Figure 2). There was no significant association between step count and hazard of CVD in the low genetic risk group, while in the moderate and high genetic risk groups, the hazard of CVD was significantly higher in the low step count compared to the high step count group (HR 1.24; 95% CI 1.10-1.40 and HR 1.37; 1.14-1.65, respectively).

### Association of PRS and continuous step count with incident CVD

Examining step count as a continuous variable using restricted cubic splines revealed more granular information on the association of step count with incidence CVD (Figure 3). There was an inverse dose-response association between the hazard of CVD and step counts up to approximately 10,000 daily steps observed in moderate and high genetic risk groups, which then plateaued. No such association was found in the low genetic risk group.

**Figure 3.**
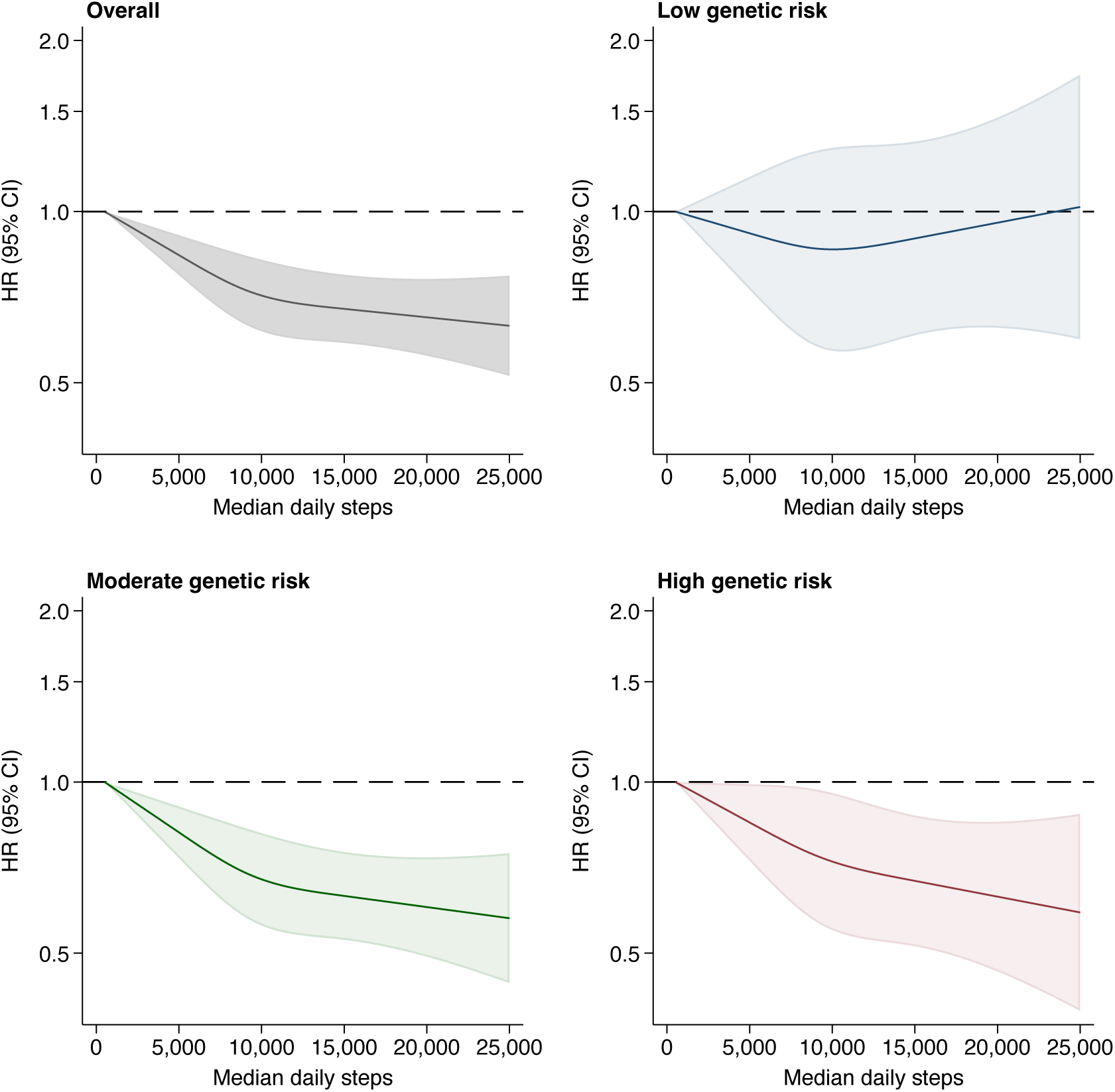
Association of median daily steps as continuous variable with incident cardiovascular disease overall and stratified by genetic risk groups. The median daily step count was modelled using restricted cubic splines with knots at the 10^th^, 50^th^, and 90^th^ percentile of the distribution of step count. The hazard ratio (solid line) and 95% CI (shaded area) are from fully adjusted Cox models.

### Association of PRS and peak 30-minute cadence with incident CVD

The 30-minute peak cadence, which represented an individual’s step intensity, was also examined in fully adjusted Cox models that did not include step count, as the two variables were strongly correlated (correlation coefficient=0.672), and the inclusion of step count did not significantly improve the models (LR test p=0.735). Lower step intensity was significantly associated with higher hazard of CVD among those at moderate genetic risk, while this trend was not statistically significant in the low and high genetic risk groups (Figure 4).

**Figure 4.**
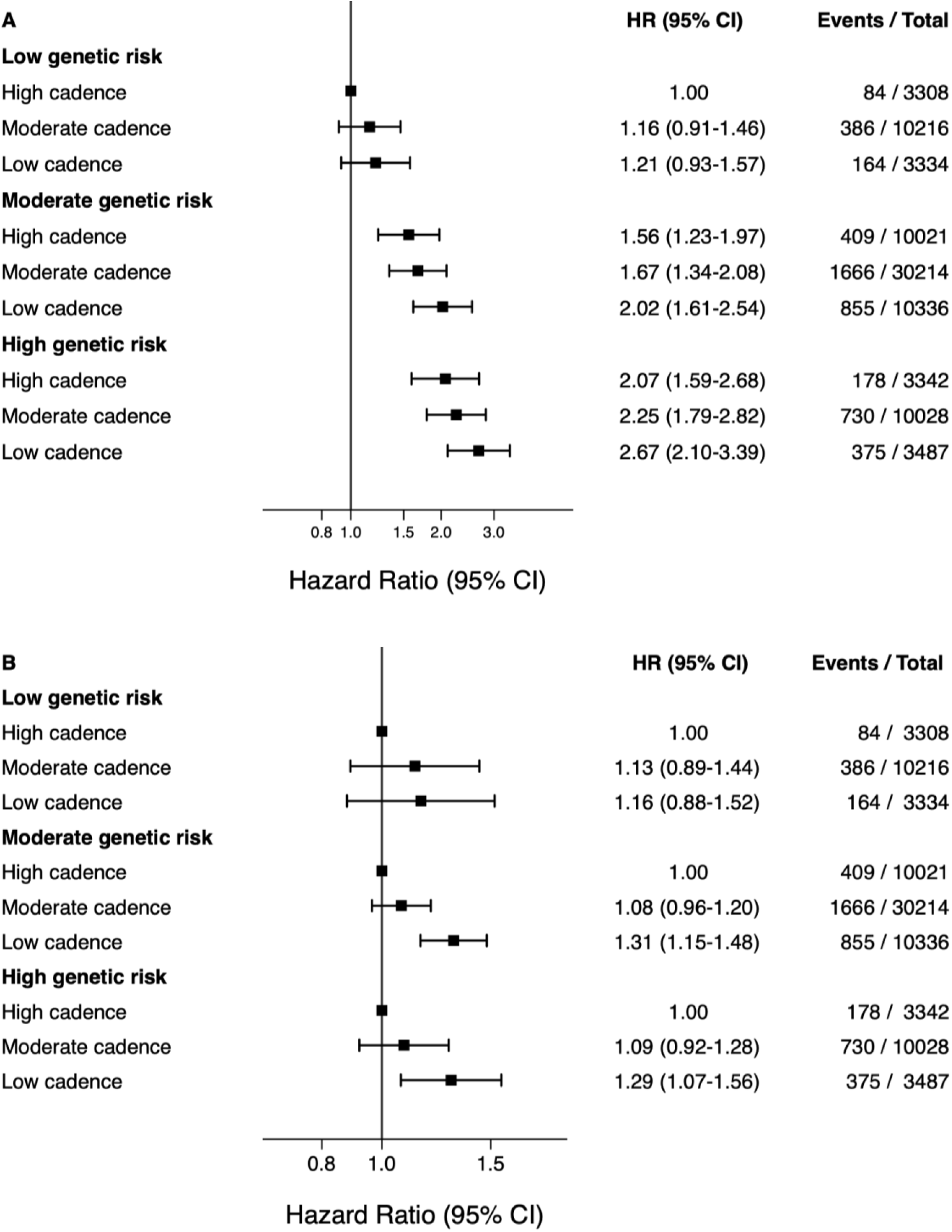
Association of genetic risk and peak 30-minute cadence with incident cardiovascular disease overall (A) and stratified by genetic risk (B). Squares represent hazard ratios (HR) and the vertical lines 95% confidence intervals

### Sensitivity analyses

Excluding events that happened in the first two years of follow-up (n=1,052) attenuated the associations of PRS and step count with incident CVD (Supplementary Figure 1). These were attenuated more considerably in the moderate genetic risk group when events in the first four years of follow-up were excluded (Supplementary Figure 2). After participants with chronic conditions or disabilities at baseline were excluded leaving 41,953 participants in the analysis (n=3,137 CVD events), we did not observe a notable change in the hazard ratios overall or in the models stratified by genetic risk (Supplementary Figure 3). Finally, when the outcomes of coronary artery disease (n=3,746) and ischaemic stroke (n=1,101) were examined separately, the effect sizes were similar for the low and moderate genetic risk groups, while in the high genetic risk group the association with step count was stronger for CAD compared to ischaemic stroke (Supplementary Figure 4, Supplementary Table 4).

## Discussion

This large prospective cohort study of 84,286 UKB participants found that low step count was associated with higher incidence of CVD, especially in individuals at moderate or high genetic risk of cardiovascular disease, after adjustment for various sociodemographic and lifestyle factors. This association was more notable below 10,000 steps/day, while there was no marked change in CVD hazard above 10,000 steps/day. Slow walking pace in the form of 30-minute cadence displayed similar associations with higher incidence of CVD.

A novel finding of this study is the difference in associations of step count with CVD between genetic risk groups, which indicates that walking may be more important for primary prevention of CVD among individuals at high genetic risk compared to low. A similar difference in the magnitude of association of PA with CVD was found in a study of self-reported walking pace,^11^ but not in other studies of self-reported PA, which demonstrated similar associations across all genetic groups.^10,29^ The significant association between PA and incident CVD found in those with high genetic risk indicates that exercise has a notable impact on CVD risk in high-risk individuals. It is also possible that longer follow-up could reveal similar associations in the low genetic risk group, as the effects of genetic predisposition might become outweighed by non-genetic factors. These findings underscore the importance of lifestyle choices to mitigate the genetic risk of CVD. Additionally, as in other studies, no multiplicative interaction between PRS and physical activity was found apart from their additive effects.^7,16,17,25^ Further comparisons with the literature are difficult, as no previous studies have examined the association between step count and genetic risk on CVD.

While in agreement with existing literature,^18,30,31^ our study found a weaker association between step count and CVD, with 24% higher risk of CVD in low vs. high step count groups in fully adjusted models, compared to 138% higher risk reported in a meta-analysis of 4 studies.^18^ A possible reason for this is that we accounted for genetic risk of CVD in our models, which may have played an important role given the strong association between genetic risk and incident CVD. Additionally, we used a stricter definition of incident CVD, focusing on coronary artery disease and ischaemic stroke, compared to other studies.^30^ Finally, we accounted for an extensive range of lifestyle factors, such as diet and sleep, which could have attenuated the observed association of step count with CVD.

The association of walking with low cardiovascular risk may be due to its impact on cardiopulmonary, circulatory, and immune functions, and its indirect effects on obesity, blood pressure, sleep quality and mental well-being.^32^ However, as in many physiological processes, after a certain threshold an individual may be in an optimal health state beyond which no further improvements can be made, resulting in the plateau we observed. Regarding peak 30-minute cadence, another UK Biobank study demonstrated an association with CVD incidence that was independent from the total daily steps,^30^ similar to our findings. Relevant literature is otherwise limited, because objectively measured walking pace has only been explored in relation to all-cause mortality.^33,34^

### Strengths and limitations

This study has various strengths, such as its prospective design, the large cohort size, and the moderately long follow-up of eight years. Additionally, the step count was recorded objectively through accelerometers, which provide a more accurate measurement of physical activity compared to self-reporting via questionnaires.^15,29,35^ However, it also has some limitations. Firstly, it is not possible to infer causal associations due to the observational study design. Similarly, individuals taking fewer steps may have done so due to preclinical CVD limiting their mobility, therefore reverse causality cannot be excluded but the risk is low based on the conducted sensitivity analyses. Secondly, even though the analyses were adjusted for multiple sociodemographic and other baseline factors, there is still the possibility of residual confounding from factors not captured in the UK Biobank. Additionally, step count data and covariate information were collected at a single time point, which for the baseline variables was a few years prior to the accelerometer study, therefore changes over time would not have been captured and adjusted for. Future studies may benefit from longer follow-up and longitudinal capture of physical activity at various time points. Furthermore, the polygenic risk score for CVD in the UKB was primarily derived from studies including individuals of European ancestry and its absolute performance was lower in non-European ancestries.^23^ Consequently, the findings of our study may not necessarily apply to individuals of different ethnic origins and additional genetic and accelerometry studies in non-European populations would add to the existing evidence. Participants were also of higher socioeconomic status than the general UK population, and due to the voluntary recruitment and low response rate of the UKB, there is a risk of healthy volunteer bias, which could lead to systematic differences between the study population and the overall UK population.^36^ Despite this source of bias, associations between risk factors and outcomes in the UKB in general and on this topic specifically were largely comparable to existing literature^37^.

### Conclusions

Overall, higher median daily step count was significantly associated with lower risk of CVD in moderate and high genetic risk individuals. Given the global prevalence of physical inactivity of approximately 31%^38^ and the increasing incidence of CVD in the ageing population, walking should be promoted as part of a wider set of prevention strategies to reduce CVD risk through the development of public health policies, as it is a low-cost, and does not require equipment or facilities.

## Data Availability

Researchers can access individual participant data by registering to the UK Biobank through https://www.ukbiobank.ac.uk/. Data fields used in the study are provided in the supplementary material.

## Acknowledgements

We would like to thank the participants of the UK Biobank, the research teams involved in the data collection at baseline and during the accelerometry study, and the researchers that developed the polygenic risk score and the step count algorithm used in this study.

