## Supplemental Material for "Joint association of genetic risk and accelerometer-based step count with cardiovascular disease: a UK-Biobank cohort study"

**Supplementary Table 1. Definition of disease diagnosis in UK Biobank**

| Condition | Classification | Codes |
|---|---|---|
| **Outcome variables** | | |
| Coronary artery disease (CAD) | ICD-9 | 410, 411, 412, 413, 414 |
| | ICD-10 | I20, I21, I22, I23, I24, I25 |
| | OPCS-4 | K40-K46, K47.1 (coronary artery bypass/endarterectomy)<br>K49, K50, K75 (transluminal coronary angioplasty) |
| | Self-report | Field 20002: 1074 (Angina), 1075 (Heart attack)<br>Field 6150: 1 (Heart attack), 2 (Angina), 3<br>Field 20004: 1070 (Coronary angioplasty), 1095 (coronary artery bypass graft), 1523 (triple heart bypass) |
| Ischaemic stroke (IS) | ICD-9 | 433, 434, 435, 436, 437, 438 |
| | ICD-10 | I63, I64, G45 |
| | OPCS-4 | L29.4, L29.5, L31.1, L31.3, L31.4 (carotid artery operations) |
| | Self-report | Field 20002: 1082 (transient ischaemic attack), 1583 (ischaemic stroke)<br>Field 6150: 3 (stroke)<br>Field 20004: 1105 (carotid artery surgery), 1109 (carotid artery angioplasty/stent) |
| **Conditions used in sensitivity analysis** | | |
| Chronic lower respiratory diseases | ICD-9 | 490-496 |
| | ICD-10 | J40-J47 |
| | Self-report | Chronic obstructive pulmonary disease, emphysema, bronchiectasis, interstitial lung disease, respiratory failure |
| Cardiovascular diseases (other than CAD and IS) | ICD-9 | 390-398, 415-432, 439-459 |
| | ICD-10 | I00-I09, I26-I28, I30-I62, I65-I99 |
| | Self-report | Cardiac problem, heart failure, leg claudication |
| Neurological conditions | ICD-9 | 330-344 |
| | ICD-10 | G10-G37, G70-G99 |
| | Self-report | Cerebral palsy, myasthenia gravis, paraplegia |
| Musculoskeletal and connective tissue disorders | ICD-9 | 710-739 |
| | ICD-10 | M00-M99 |
| | Self-report | Osteoarthritis, rheumatoid arthritis |
| Cancer | ICD-9 | 140-209 |
| | ICD-10 | C00-C97 |
| Abnormalities of gait and mobility | ICD-10 | R26-R29 |

*ICD: International Classification of Diseases and Related Health Problems*

*OPCS: Office of Population Censuses and Surveys Classification of Surgical Operations*

**Supplementary Table 2. Field ID in the UK Biobank for the variables included in the analysis**

| Variable | Field ID |
|---|---|
| Month / Year of birth | 52 / 34 |
| End time of accelerometer wear | 90011 |
| Date lost to follow-up | 191 |
| Hospital Episode Statistics inpatient dataset (diagnoses, operations) | Category 2000 |
| Death register (date of death, cause of death) | Category 100093 |
| Acceleration data* | 90001 |
| Standard PRS for cardiovascular disease (CVD) | 26223 |
| Sex | 31 |
| Ethnicity | 21000 |
| Education | 6138 |
| Employment status | 6142, 20119 |
| Townsend Deprivation Index | 22189 |
| Smoking status | 20116 |
| Alcohol consumption | 1558, 20117 |
| Fruit and vegetable consumption | 1289, 1299, 1309 |
| Oily and non-oily fish consumption | 1329, 1339 |
| Red and processed meat consumption | 1349, 1369, 1379, 1389 |
| Salt consumption | 1478 |
| Accelerometry-derived sleep duration | 40046 |
| Family history of cardiovascular disease | 20107, 20110, 20111 |
| Total cholesterol | 30690 |
| Glucose | 30740 |
| Glycated Haemoglobin (HbA1c) | 30750 |
| Systolic blood pressure | 4080 (if missing 93) |
| Diastolic blood pressure | 4079 (if missing 94) |
| Body mass index | 21001 |
| Self-reported illnesses | 20002 |
| Self-reported operations | 20004 |
| Medications | 20003, 6177, 6153 |
| Doctor diagnosed conditions | 2443, 6150 |

** Accelerometry-derived step count and cadence were derived from that variable through work of this project's team and are not publicly available in the UK Biobank.*

**Supplementary Table 3. Baseline characteristics of study cohort by genetic risk group**

| | Genetic risk | | | | |
|---|---|---|---|---|---|
| | Low (N=16,858) | Moderate (N=50,571) | High (N=16,857) | Total (N=84,286) | p-value |
| **Age (years)** | 63·4 (56·2-68·5) | 63·0 (55·8-68·2) | 62·4 (55·6-67·8) | 62·9 (55·8-68·2) | <0·001 |
| **Reported sex** | | | | | <0·001 |
| Female | 9,685 (57·5) | 29,221 (57·8) | 10,088 (59·8) | 48,994 (58·1) | |
| Male | 7,173 (42·5) | 21,350 (42·2) | 6,769 (40·2) | 35,292 (41·9) | |
| **Ethnicity** | | | | | 0·003 |
| White | 16,363 (97·1) | 49,162 (97·2) | 16,301 (96·7) | 81,826 (97·1) | |
| Non-white | 495 (2·9) | 1,409 (2·8) | 556 (3·3) | 2,460 (2·9) | |
| **Educational attainment** | | | | | <0·001 |
| Basic education | 3,568 (21·2) | 11,159 (22·1) | 4,015 (23·8) | 18,742 (22·2) | |
| Further education | 5,477 (32·5) | 16,863 (33·3) | 5,641 (33·5) | 27,981 (33·2) | |
| University degree | 7,813 (46·3) | 22,549 (44·6) | 7,201 (42·7) | 37,563 (44·6) | |
| **Employment status** | | | | | <0·001 |
| Employed | 10,549 (62·6) | 32,074 (63·4) | 10,939 (64·9) | 53,562 (63·5) | |
| Not employed | 1,113 (6·6) | 3,365 (6·7) | 1,176 (7·0) | 5,654 (6·7) | |
| Retired | 5,196 (30·8) | 15,132 (29·9) | 4,742 (28·1) | 25,070 (29·7) | |
| **Townsend Deprivation Index** | | | | | 0·254 |
| Least deprived | 8,619 (51·1) | 25,842 (51·1) | 8,468 (50·2) | 42,929 (50·9) | |
| 2nd Quintile | 3,827 (22·7) | 11,363 (22·5) | 3,832 (22·7) | 19,022 (22·6) | |
| 3rd Quintile | 2,362 (14·0) | 6,932 (13·7) | 2,396 (14·2) | 11,690 (13·9) | |
| 4th Quintile | 1,543 (9·2) | 4,798 (9·5) | 1,595 (9·5) | 7,936 (9·4) | |
| Most deprived | 507 (3·0) | 1,636 (3·2) | 566 (3·4) | 2,709 (3·2) | |
| **Country** | | | | | <0·001 |
| England | 15,198 (90·2) | 45,392 (89·8) | 15,028 (89·1) | 75,618 (89·7) | |
| Scotland | 980 (5·8) | 3,308 (6·5) | 1,183 (7·0) | 5,471 (6·5) | |
| Wales | 680 (4·0) | 1,871 (3·7) | 646 (3·8) | 3,197 (3·8) | |
| **Smoking status** | | | | | 0·041 |
| Never | 9,934 (58·9) | 29,387 (58·1) | 9,739 (57·8) | 49,060 (58·2) | |
| Previous | 5,867 (34·8) | 17,753 (35·1) | 5,938 (35·2) | 29,558 (35·1) | |
| Current | 1,057 (6·3) | 3,431 (6·8) | 1,180 (7·0) | 5,668 (6·7) | |
| **Alcohol consumption** | | | | | 0·029 |
| Daily | 3,969 (23·5) | 11,617 (23·0) | 3,786 (22·5) | 19,372 (23·0) | |
| 1-4 /week | 8,638 (51·2) | 26,173 (51·8) | 8,621 (51·1) | 43,432 (51·5) | |
| < 1/week | 3,328 (19·7) | 10,105 (20·0) | 3,529 (20·9) | 16,962 (20·1) | |
| Never | 923 (5·5) | 2,676 (5·3) | 921 (5·5) | 4,520 (5·4) | |
| **Red/processed meat** | | | | | 0·038 |
| <1 time/week | 1,364 (8·1) | 4,026 (8·0) | 1,382 (8·2) | 6,772 (8·0) | |
| 1-3 times/week | 6,437 (38·2) | 19,577 (38·7) | 6,556 (38·9) | 32,570 (38·6) | |
| 3-4 times/week | 4,847 (28·8) | 14,874 (29·4) | 4,968 (29·5) | 24,689 (29·3) | |
| 5+ times/week | 4,210 (25·0) | 12,094 (23·9) | 3,951 (23·4) | 20,255 (24·0) | |
| **Fish consumption** | | | | | 0·012 |

| | | | | | |
|---|---|---|---|---|---|
| 0-1 times/week | 8,368 (49·6) | 24,622 (48·7) | 8,098 (48·0) | 41,088 (48·7) | |
| 2+ times/week | 8,490 (50·4) | 25,949 (51·3) | 8,759 (52·0) | 43,198 (51·3) | |
| **Fruit/veg consumption** | | | | | 0·089 |
| <5 servings/day | 3,510 (20·8) | 10,290 (20·3) | 3,435 (20·4) | 17,235 (20·4) | |
| 5-7·9 servings/day | 7,229 (42·9) | 21,312 (42·1) | 7,149 (42·4) | 35,690 (42·3) | |
| 8+ servings/day | 6,119 (36·3) | 18,969 (37·5) | 6,273 (37·2) | 31,361 (37·2) | |
| **Salt consumption** | | | | | 0·018 |
| Never/rarely | 10,021 (59·4) | 30,431 (60·2) | 10,257 (60·8) | 50,709 (60·2) | |
| Sometimes | 4,525 (26·8) | 13,333 (26·4) | 4,465 (26·5) | 22,323 (26·5) | |
| Usually/Always | 2,312 (13·7) | 6,807 (13·5) | 2,135 (12·7) | 11,254 (13·4) | |
| **Sleep duration** | | | | | 0·086 |
| 6h or fewer | 5,749 (34·1) | 17,181 (34·0) | 5,733 (34·0) | 28,663 (34·0) | |
| 7-8h | 10,728 (63·6) | 32,204 (63·7) | 10,671 (63·3) | 53,603 (63·6) | |
| 9h or more | 381 (2·3) | 1,186 (2·3) | 453 (2·7) | 2,020 (2·4) | |
| **Family history of CVD** | 8,407 (49·9) | 28,426 (56·2) | 10,477 (62·2) | 47,310 (56·1) | <0·001 |
| **Diabetes** | 831 (4·9) | 2,806 (5·5) | 989 (5·9) | 4,626 (5·5) | <0·001 |
| **Hypercholesterolaemia** | 12,837 (76·1) | 40,273 (79·6) | 13,801 (81·9) | 66,911 (79·4) | <0·001 |
| **Hypertension** | 7,271 (43·1) | 24,638 (48·7) | 9,183 (54·5) | 41,092 (48·8) | <0·001 |
| **Body Mass Index (kg/m$^2$)** | 25·7 (23·3-28·6) | 25·9 (23·5-28·8) | 26·0 (23·6-29·0) | 25·9 (23·5-28·8) | <0·001 |
| **Systolic blood pressure (mmHg)** | 134·3 ± 17·8 | 136·2 ± 18·0 | 138·1 ± 18·5 | 136·2 ± 18·1 | <0·001 |
| **Diastolic blood pressure (mmHg)** | 80·6 ± 9·9 | 81·7 ± 10·0 | 82·8 ± 9·9 | 81·7 ± 10·0 | <0·001 |
| **Median daily steps** | 9,192 (6,991-11,794) | 9,127 (6,897-11,748) | 9,092 (6,872-11,699) | 9,134 (6,910-11,749) | 0·099 |
| **Median daily step count** | | | | | 0·023 |
| Low | 3,401 (20·2) | 10,646 (21·1) | 3,620 (21·5) | 17,667 (21·0) | |
| Moderate | 10,074 (59·8) | 29,885 (59·1) | 9,989 (59·3) | 49,948 (59·3) | |
| High | 3,383 (20·1) | 10,040 (19·9) | 3,248 (19·3) | 16,671 (19·8) | |
| **Peak 30-min cadence (steps/min)** | 93·2 ± 16·1 | 93·0 ± 16·3 | 92·9 ± 16·4 | 93·0 ± 16·3 | 0·226 |
| **Season of wear** | | | | | 0·614 |
| Autumn | 4,962 (29·4) | 14,904 (29·5) | 5,010 (29·7) | 24,876 (29·5) | |
| Winter | 3,608 (21·4) | 10,600 (21·0) | 3,499 (20·8) | 17,707 (21·0) | |
| Spring | 3,824 (22·7) | 11,730 (23·2) | 3,846 (22·8) | 19,400 (23·0) | |
| Summer | 4,464 (26·5) | 13,337 (26·4) | 4,502 (26·7) | 22,303 (26·5) | |

*Summary statistics are presented as n (%) for categorical variables, mean ± SD for continuous normally distributed, and median (IQR) for not normally distributed variables. p-values derive from Pearson's chi-squared test for categorical variables and linear regression for continuous variables. CVD: Cardiovascular disease*

**Supplementary Table 4. Association of genetic risk and median daily step count with incident cardiovascular disease components overall and from stratified models**

| | Coronary artery disease (CAD) | | Ischaemic stroke | |
|---|---|---|---|---|
| | **Events/Total** | **HR (95% CI)** | **Events/Total** | **HR (95% CI)** |
| *Low genetic risk* | | | | |
| High step count | 88/3,383 | 1·00 | 28/3,383 | 1·00 |
| Moderate step count | 262/10,074 | 0·93 (0·73-1·19) | 103/10,074 | 1·13 (0·75-1·72) |
| Low step count | 116/3,401 | 1·03 (0·78-1·36) | 37/3,401 | 1·06 (0·65-1·74) |
| *Moderate genetic risk* | | | | |
| High step count | 361/10,040 | 1·35 (1·07-1·71) | 110/10,040 | 1·32 (0·87-2·01) |
| Moderate step count | 1,284/29,885 | 1·51 (1·22-1·88) | 393/29,885 | 1·46 (0·99-2·14) |
| Low step count | 597/10,646 | 1·65 (1·32-2·07) | 185/10,646 | 1·68 (1·13-2·52) |
| *High genetic risk* | | | | |
| High step count | 152/3,248 | 1·78 (1·36-2·31) | 39/3,248 | 1·50 (0·92-2·43) |
| Moderate step count | 590/9,989 | 2·11 (1·69-2·64) | 135/9,989 | 1·52 (1·01-2·29) |
| Low step count | 296/3,620 | 2·42 (1·90-3·08) | 71/3,620 | 1·93 (1·24-3·00) |
| P (association) | | <0·001 | | 0·002 |
| **Stratified models** | | | | |
| *Low genetic risk* | | | | |
| High step count | 88/3,383 | 1·00 | 28/3,383 | 1·00 |
| Moderate step count | 262/10,074 | 0·93 (0·72-1·18) | 103/10,074 | 1·14 (0·74-1·73) |
| Low step count | 116/3,401 | 0·99 (0·74-1·32) | 37/3,401 | 1·07 (0·64-1·78) |
| P (association) | | 0·757 | | 0·826 |
| *Moderate genetic risk* | | | | |
| High step count | 361/10,040 | 1·00 | 110/10,040 | 1·00 |
| Moderate step count | 1,284/29,885 | 1·12 (0·99-1·26) | 393/29,885 | 1·10 (0·89-1·36) |
| Low step count | 597/10,646 | 1·23 (1·07-1·41) | 185/10,646 | 1·27 (0·99-1·63) |
| P (association) | | 0·012 | | 0·127 |
| *High genetic risk* | | | | |
| High step count | 152/3,248 | 1·00 | 39/3,248 | 1·00 |
| Moderate step count | 590/9,989 | 1·20 (1·01-1·44) | 135/9,989 | 1·02 (0·71-1·46) |
| Low step count | 296/3,620 | 1·38 (1·12-1·69) | 71/3,620 | 1·30 (0·86-1·95) |
| P (association) | | 0·008 | | 0·239 |

*Hazard Ratios and 95% confidence intervals derive from fully adjusted Cox models using age as the time scale. p-values derive from a Wald test of overall association.*

**Supplementary Figure 1. Sensitivity analysis excluding cardiovascular events in the first two years of follow-up overall (A) and stratified by genetic risk (B)**

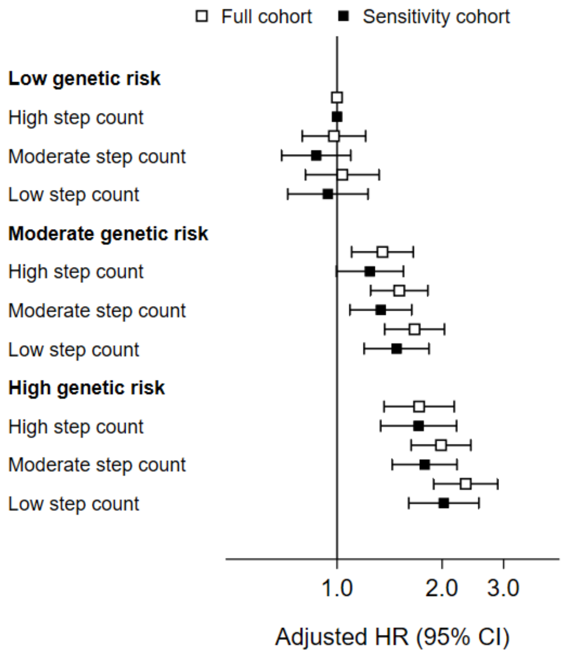


| | Full cohort | | Sensitivity cohort | |
|---|---|---|---|---|
| | Event / Total | HR (95% CI) | Event / Total | HR (95% CI) |
| **Low genetic risk** | | | | |
| High step count | 116 / 3383 | 1.00 | 100 / 3367 | 1.00 |
| Moderate step count | 365 / 10074 | 0.98 (0.80-1.21) | 288 / 9997 | 0.87 (0.69-1.09) |
| Low step count | 153 / 3401 | 1.04 (0.81-1.32) | 125 / 3373 | 0.94 (0.72-1.23) |
| **Moderate genetic risk** | | | | |
| High step count | 471 / 10040 | 1.35 (1.10-1.65) | 371 / 9940 | 1.24 (1.00-1.55) |
| Moderate step count | 1677 / 29885 | 1.51 (1.25-1.82) | 1297 / 29505 | 1.34 (1.09-1.64) |
| Low step count | 782 / 10646 | 1.67 (1.37-2.03) | 616 / 10480 | 1.48 (1.20-1.83) |
| **High genetic risk** | | | | |
| High step count | 191 / 3248 | 1.72 (1.36-2.17) | 160 / 3217 | 1.71 (1.33-2.20) |
| Moderate step count | 725 / 9989 | 1.99 (1.63-2.42) | 560 / 9824 | 1.78 (1.44-2.21) |
| Low step count | 367 / 3620 | 2.34 (1.89-2.89) | 278 / 3531 | 2.02 (1.61-2.55) |

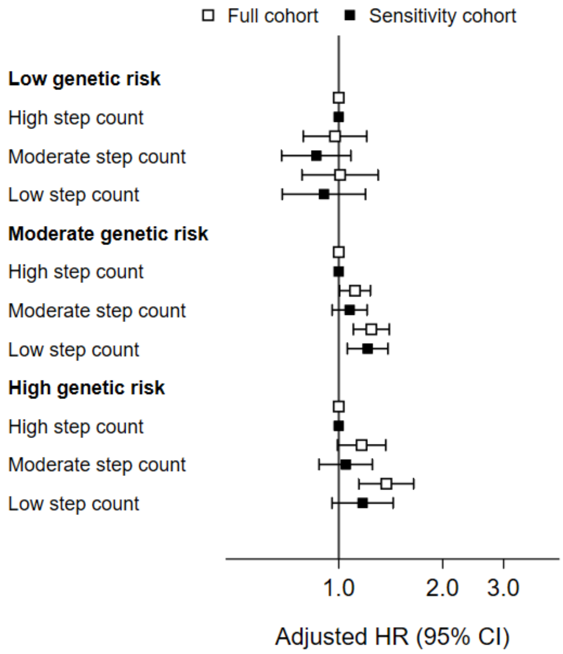


| | Full cohort | | Sensitivity cohort | |
|---|---|---|---|---|
| | Event / Total | HR (95% CI) | Event / Total | HR (95% CI) |
| **Low genetic risk** | | | | |
| High step count | 116 / 3383 | 1.00 | 100 / 3367 | 1.00 |
| Moderate step count | 365 / 10074 | 0.98 (0.79-1.21) | 288 / 9997 | 0.86 (0.68-1.08) |
| Low step count | 153 / 3401 | 1.01 (0.78-1.30) | 125 / 3373 | 0.91 (0.69-1.20) |
| **Moderate genetic risk** | | | | |
| High step count | 471 / 10040 | 1.00 | 371 / 9940 | 1.00 |
| Moderate step count | 1677 / 29885 | 1.11 (1.00-1.24) | 1297 / 29505 | 1.08 (0.96-1.21) |
| Low step count | 782 / 10646 | 1.24 (1.10-1.40) | 616 / 10480 | 1.21 (1.06-1.39) |
| **High genetic risk** | | | | |
| High step count | 191 / 3248 | 1.00 | 160 / 3217 | 1.00 |
| Moderate step count | 725 / 9989 | 1.16 (0.99-1.37) | 560 / 9824 | 1.05 (0.88-1.25) |
| Low step count | 367 / 3620 | 1.37 (1.14-1.65) | 278 / 3531 | 1.17 (0.96-1.44) |

*Squares represent hazard ratios (HR) and the vertical lines 95% confidence intervals (95% CI) from fully adjusted Cox models using age as time scale.*

**Supplementary Figure 2. Sensitivity analysis excluding cardiovascular events in the first four years of follow-up overall (A) and stratified by genetic risk (B)**

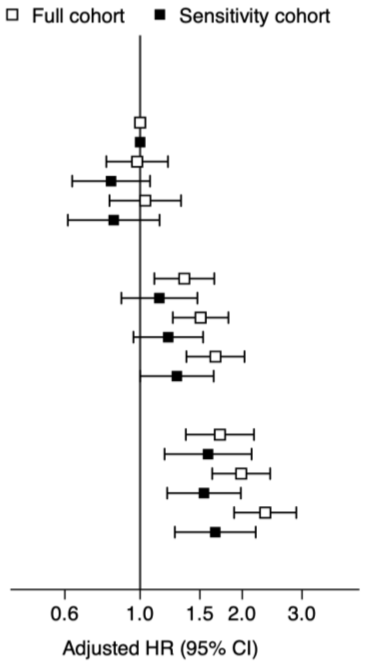


| A | Full cohort | | Sensitivity cohort | |
|---|---|---|---|---|
| **Low genetic risk** | **Event / Total** | **HR (95% CI)** | **Event / Total** | **HR (95% CI)** |
| High step count | 116 / 3383 | 1.00 | 75 / 3342 | 1.00 |
| Moderate step count | 365 / 10074 | 0.98 (0.80-1.21) | 210 / 9919 | 0.82 (0.63-1.07) |
| Low step count | 153 / 3401 | 1.04 (0.81-1.32) | 87 / 3335 | 0.84 (0.61-1.14) |
| **Moderate genetic risk** | | | | |
| High step count | 471 / 10040 | 1.35 (1.10-1.65) | 253 / 9822 | 1.14 (0.88-1.47) |
| Moderate step count | 1677 / 29885 | 1.51 (1.25-1.82) | 891 / 29099 | 1.21 (0.96-1.53) |
| Low step count | 782 / 10646 | 1.67 (1.37-2.03) | 411 / 10275 | 1.28 (1.00-1.65) |
| **High genetic risk** | | | | |
| High step count | 191 / 3248 | 1.72 (1.36-2.17) | 108 / 3165 | 1.59 (1.18-2.13) |
| Moderate step count | 725 / 9989 | 1.99 (1.63-2.42) | 360 / 9624 | 1.54 (1.20-1.98) |
| Low step count | 367 / 3620 | 2.34 (1.89-2.89) | 174 / 3427 | 1.67 (1.27-2.19) |

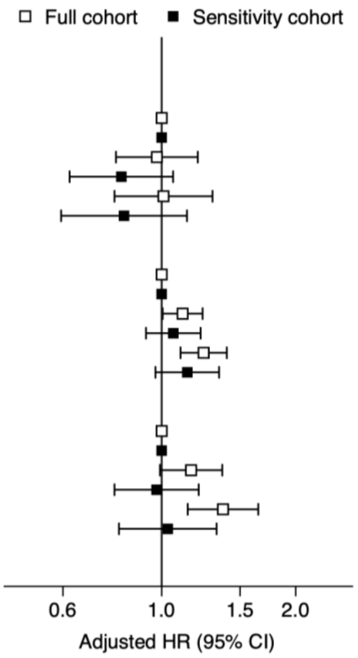


| B | Full cohort | | Sensitivity cohort | |
|---|---|---|---|---|
| **Low genetic risk** | **Event / Total** | **HR (95% CI)** | **Event / Total** | **HR (95% CI)** |
| High step count | 116 / 3383 | 1.00 | 75 / 3342 | 1.00 |
| Moderate step count | 365 / 10074 | 0.98 (0.79-1.21) | 210 / 9919 | 0.81 (0.62-1.06) |
| Low step count | 153 / 3401 | 1.01 (0.78-1.30) | 87 / 3335 | 0.82 (0.59-1.14) |
| **Moderate genetic risk** | | | | |
| High step count | 471 / 10040 | 1.00 | 253 / 9822 | 1.00 |
| Moderate step count | 1677 / 29885 | 1.11 (1.00-1.24) | 891 / 29099 | 1.06 (0.92-1.22) |
| Low step count | 782 / 10646 | 1.24 (1.10-1.40) | 411 / 10275 | 1.14 (0.97-1.35) |
| **High genetic risk** | | | | |
| High step count | 191 / 3248 | 1.00 | 108 / 3165 | 1.00 |
| Moderate step count | 725 / 9989 | 1.16 (0.99-1.37) | 360 / 9624 | 0.97 (0.78-1.21) |
| Low step count | 367 / 3620 | 1.37 (1.14-1.65) | 174 / 3427 | 1.03 (0.80-1.33) |

*Squares represent hazard ratios (HR) and the vertical lines 95% confidence intervals (95% CI) from fully adjusted Cox models using age as time scale.*

**Supplementary Figure 3. Sensitivity analysis excluding individuals with comorbid conditions at the time of accelerometer wear overall (A) and stratified by genetic risk (B)**

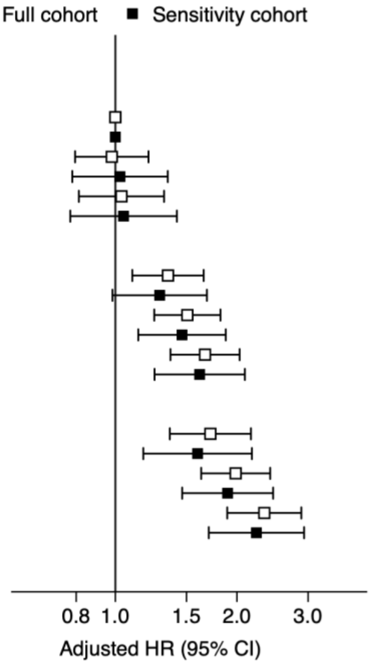


**A**

| | Full cohort | | Disease-free cohort | |
|---|---|---|---|---|
| | **Event / Total** | **HR (95% CI)** | **Event / Total** | **HR (95% CI)** |
| **Low genetic risk** | | | | |
| High step count | 116 / 3383 | 1.00 | 66 / 1437 | 1.00 |
| Moderate step count | 365 / 10074 | 0.98 (0.80-1.21) | 246 / 4908 | 1.03 (0.78-1.35) |
| Low step count | 153 / 3401 | 1.04 (0.81-1.32) | 117 / 1948 | 1.05 (0.77-1.42) |
| **Moderate genetic risk** | | | | |
| High step count | 471 / 10040 | 1.35 (1.10-1.65) | 273 / 4554 | 1.29 (0.98-1.69) |
| Moderate step count | 1677 / 29885 | 1.51 (1.25-1.82) | 1035 / 14324 | 1.46 (1.14-1.88) |
| Low step count | 782 / 10646 | 1.67 (1.37-2.03) | 575 / 6219 | 1.62 (1.25-2.09) |
| **High genetic risk** | | | | |
| High step count | 191 / 3248 | 1.72 (1.36-2.17) | 103 / 1426 | 1.60 (1.17-2.18) |
| Moderate step count | 725 / 9989 | 1.99 (1.63-2.42) | 450 / 4934 | 1.90 (1.46-2.46) |
| Low step count | 367 / 3620 | 2.34 (1.89-2.89) | 272 / 2203 | 2.23 (1.70-2.93) |

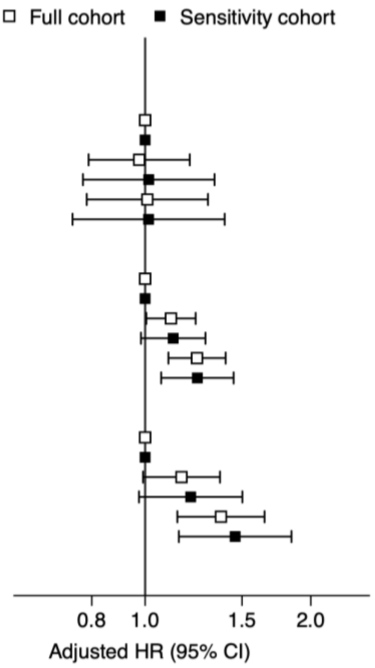


**B**

| | Full cohort | | Disease-free cohort | |
|---|---|---|---|---|
| | **Event / Total** | **HR (95% CI)** | **Event / Total** | **HR (95% CI)** |
| **Low genetic risk** | | | | |
| High step count | 116 / 3383 | 1.00 | 66 / 1437 | 1.00 |
| Moderate step count | 365 / 10074 | 0.98 (0.79-1.21) | 246 / 4908 | 1.02 (0.77-1.34) |
| Low step count | 153 / 3401 | 1.01 (0.78-1.30) | 117 / 1948 | 1.01 (0.74-1.39) |
| **Moderate genetic risk** | | | | |
| High step count | 471 / 10040 | 1.00 | 273 / 4554 | 1.00 |
| Moderate step count | 1677 / 29885 | 1.11 (1.00-1.24) | 1035 / 14324 | 1.12 (0.98-1.29) |
| Low step count | 782 / 10646 | 1.24 (1.10-1.40) | 575 / 6219 | 1.24 (1.07-1.45) |
| **High genetic risk** | | | | |
| High step count | 191 / 3248 | 1.00 | 103 / 1426 | 1.00 |
| Moderate step count | 725 / 9989 | 1.16 (0.99-1.37) | 450 / 4934 | 1.21 (0.97-1.50) |
| Low step count | 367 / 3620 | 1.37 (1.14-1.65) | 272 / 2203 | 1.46 (1.15-1.84) |

*Comorbid conditions included chronic respiratory, cardiovascular, musculoskeletal or neurological conditions and cancer based on hospital records, or self-reported disability prior to the study period. Squares represent hazard ratios (HR) and the vertical lines 95% confidence intervals (95% CI) from fully adjusted Cox models using age as time scale.*

**Supplementary Figure 4. Association of genetic risk and step count with components of cardiovascular disease (coronary artery disease and ischaemic stroke)**

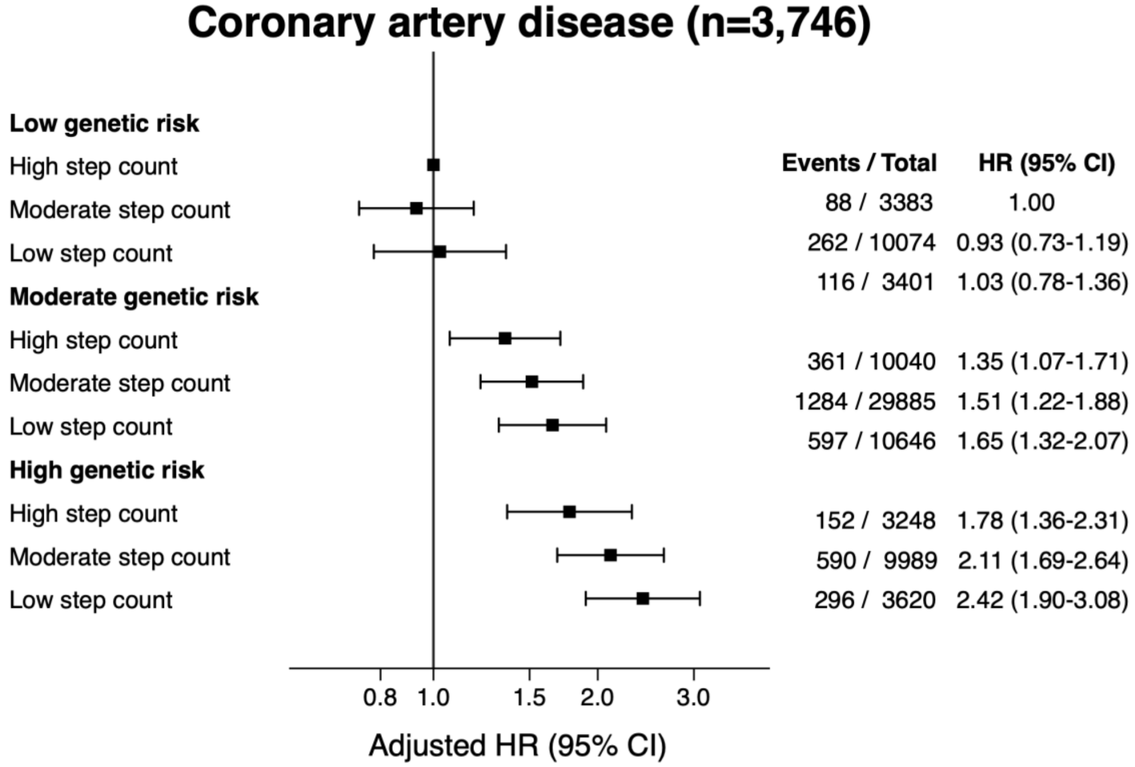


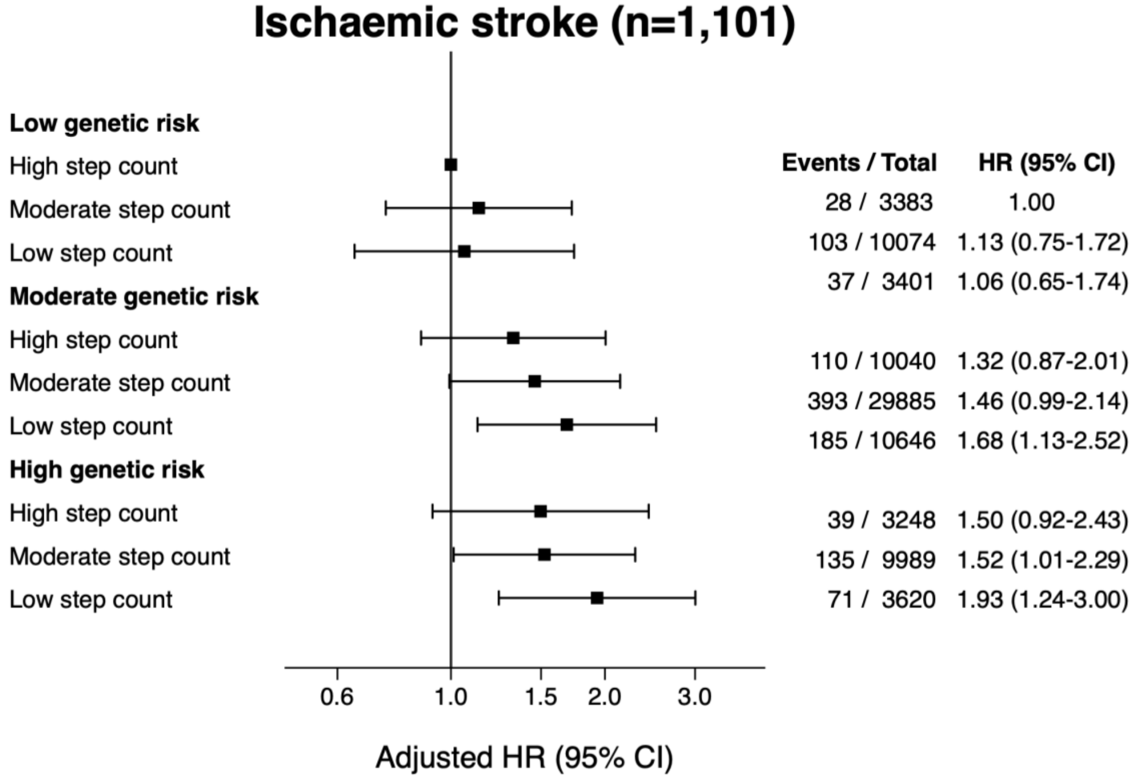


*Squares represent hazard ratios (HR) and the vertical lines 95% confidence intervals (95% CI) from fully adjusted Cox models using age as time scale.*
